# Local SARS-CoV-2 Peptide-Specific Immune Responses in Lungs of Convalescent and Uninfected Human Subjects

**DOI:** 10.1101/2021.09.02.21263042

**Authors:** Kayla F. Goliwas, Anthony M. Wood, Christopher S. Simmons, Rabisa Khan, Saad A. Khan, Yong Wang, Joel L. Berry, Mohammad Athar, James A. Mobley, Young-il Kim, Victor J. Thannickal, Kevin S. Harrod, James M. Donahue, Jessy S. Deshane

## Abstract

Multi-specific and long-lasting T cell immunity have been recognized as indicators for long term protection against pathogens including the novel coronavirus SARS-CoV-2, the causative agent of the COVID-19 pandemic. Functional significance of peripheral memory T cells in individuals recovering from COVID-19 (COVID-19^+^) are beginning to be appreciated; but little is known about lung resident memory T cells (lung TRM) in SARS-CoV-2 infection. Here, we utilize a perfused three dimensional (3D) human lung tissue model and identify pre-existing local T cell immunity against SARS-CoV-2 proteins in lung tissues. We report *ex vivo* maintenance of functional multi-specific IFN-γ secreting lung TRM in COVID-19^+^ and their induction in lung tissues of vaccinated COVID-19^+^. Importantly, we identify SARS-CoV-2 peptide-responding B cells and IgA^+^ plasma cells in lung tissues of COVID-19^+^ in *ex vivo* 3D-tissue models. Our study highlights the importance of balanced and local anti-viral immune response in the lung with persistent induction of TRM and IgA^+^ plasma cells for future protection against SARS-CoV-2 infection. Further, our data suggest that inclusion of multiple viral antigens in vaccine approaches may broaden the functional profile of memory T cells to combat the severity of coronavirus infection.

## INTRODUCTION

COVID-19, the disease caused by the novel coronavirus SARS-CoV-2, is a global health concern ^1-3^. Infected individuals develop lymphopenia and demonstrate hyperactivated and exhausted T cell responses that contribute to prolonged immune dysregulation, a hallmark of SARS-CoV-2 infection^4-8^. Although vaccine efforts have been successful, the emergence of variants^9,10^, persistence of infection and vaccine hesitancy continue to pose problems for eradication and management of COVID-19. All approved COVID-19 vaccines, thus far, utilize intramuscular delivery of SARS-CoV-2 spike mRNA or inactivated virus to elicit humoral immunity, a strategy that has not offered long-term protection.

Evidence from previous zoonotic coronaviruses indicates that an important determinant for recovery and long-term protection is coronavirus-specific T cell immunity^11-13^. During the initial phase of the pandemic, 20-50% unexposed individuals showed T cell reactivity to SARS-CoV-2 antigen peptide pools^6,7,14-17^. Pre-existing CD4 and CD8 T cell responses against both structural (nucleocapsid, N) and non-structural regions of SARS-CoV-2 were noted in peripheral blood mononuclear cells of COVID-19 convalescent individuals^6,7,14-23^. In the majority of COVID-19 recovering individuals (COVID-19^+^), larger overall SARS-CoV-2-specific CD4 and CD8 T cell responses were seen with severe disease^18,22,23^. However, an increase in polyfunctional CD8^+^ T cells were noted in mild cases^18,22,23^. Furthermore, long-lasting memory T cells displayed robust cross-reactivity to N protein of SARS-CoV and SARS-CoV-2^24,25^. Interestingly, SARS-CoV-2-specfic T cells are present in individuals with no prior history of SARS suggesting pre-existing cross-reactive immune memory to seasonal coronaviruses^7,17^.

As memory T cell response induced by previous viral pathogens can shape susceptibility to subsequent viral infections including SARS-CoV-2, and/or influence clinical severity of COVID-19, pre-existing memory T cells that recognize SARS-CoV-2 have been implicated^22,24-29^. Upon SARS-CoV-2 exposure, a faster and stronger immune response is anticipated in individuals with pre-existing T cell immunity^7^. Additionally, increased memory T follicular helper CD4^+^ T cells could facilitate a rapid SARS-CoV-2 neutralizing antibody response^30^. The increase in memory CD4^+^ and CD8^+^ T cells may enable direct antiviral immunity in the lungs and nasopharynx; pre-existing CD4^+^ T cell memory could influence vaccination outcomes, leading to a quicker robust immune response and development of neutralizing antibodies^31^. Alternatively, pre-existing immunity could be detrimental due to exuberant antibody-mediated inflammation or from inadequate immune responses^32^. Thus, accurate measurements of pre-existing T cell immunity are essential to an understanding of disease severity and effective vaccine responses. The assessment of a complete SARS-CoV-2 reactive T cell pool has been challenging; both the breadth and depth of the SARS-CoV-2 specific T cell responses are not fully understood, as circulating T cells may not represent lung-specific responses to viral infection and/or reflect direct anti-viral immunity in the lungs or the nasopharynx.

In addition to T cell immunity, the most anticipated and monitored protective response is the evolution of antibody immunity to SARS-CoV-2 ^33^. While waning antibody levels have been reported after SARS-CoV-2 infection ^34-37^, very few studies have evaluated the nature and quality of the memory B cells^33^ that would be required to produce antibodies upon reinfection. Additionally, early SARS-CoV-2–specific humoral responses were dominated by IgA and not IgG antibodies ^38,39^; but their presence and function in the lungs of COVID-19^+^ are largely unknown.

We utilized a perfused three-dimensional lung tissue culture model that maintains the tissue architecture of the human lung for the assessment of pre-existing T cell immunity in the lung tissues of previously uninfected (UN) and COVID-19^+^ individuals. Utilizing this model, we demonstrate local immune responses to SARS-CoV-2 peptide pools in the lungs of UN individuals induced by pre-existing T cell immunity and significant memory T cell response in the lungs of COVID-19^+^ individuals. Importantly, we identify, for the first time, significant IgA levels and IgA-secreting plasma cells in COVID-19 infected lungs.

## RESULTS

### *Ex Vivo* Perfusion of Human Lung Tissues

Remnant surgical specimen from individuals (no history of SARS-CoV-2 infection) undergoing lung resection surgeries were collected. For *ex vivo* culture, 5 mm diameter tissue cores were generated and one tissue core was placed into the central chamber of a bioreactor containing a mixture of extracellular matrix (ECM) components. The tissue/ECM support was penetrated with five 400 micron Teflon coated stainless steel wires to generate through-channels for adequate tissue perfusion. Wires were removed following ECM polymerization. A serum-free, defined tissue culture media was then perfused from a media reservoir through the tissue volume and collected in a collection reservoir using a peristaltic pump (**Figure 1A-B**). Using this culture system, we observed maintenance of histologic tissue architecture (**Extended Fig. 1A**), as well as cell density (cells/area, **Extended Fig. 1B**) over a two-week culture period. Further, lactate dehydrogenase (LDH) remained unchanged during culture (**Extended Fig. 1C**), indicating sustained viability. The ECM composition utilized here has been used with earlier prototype bioreactors to generate viable cell culture models of lung and breast carcinoma (Goliwas et. al. 2021; Goliwas et al, 2016). These platforms were adapted here for the *ex vivo* culture of human lung tissues. Cell phenotyping showed maintenance of lung epithelial and endothelial cells, as well as fibroblasts and lymphocytes including CD8^+^ T cells (**Extended Figs. 1D-H**), and proteomic analyses showed maintenance of the lung ECM (**Extended Fig. 1I**).

**Figure 1:**
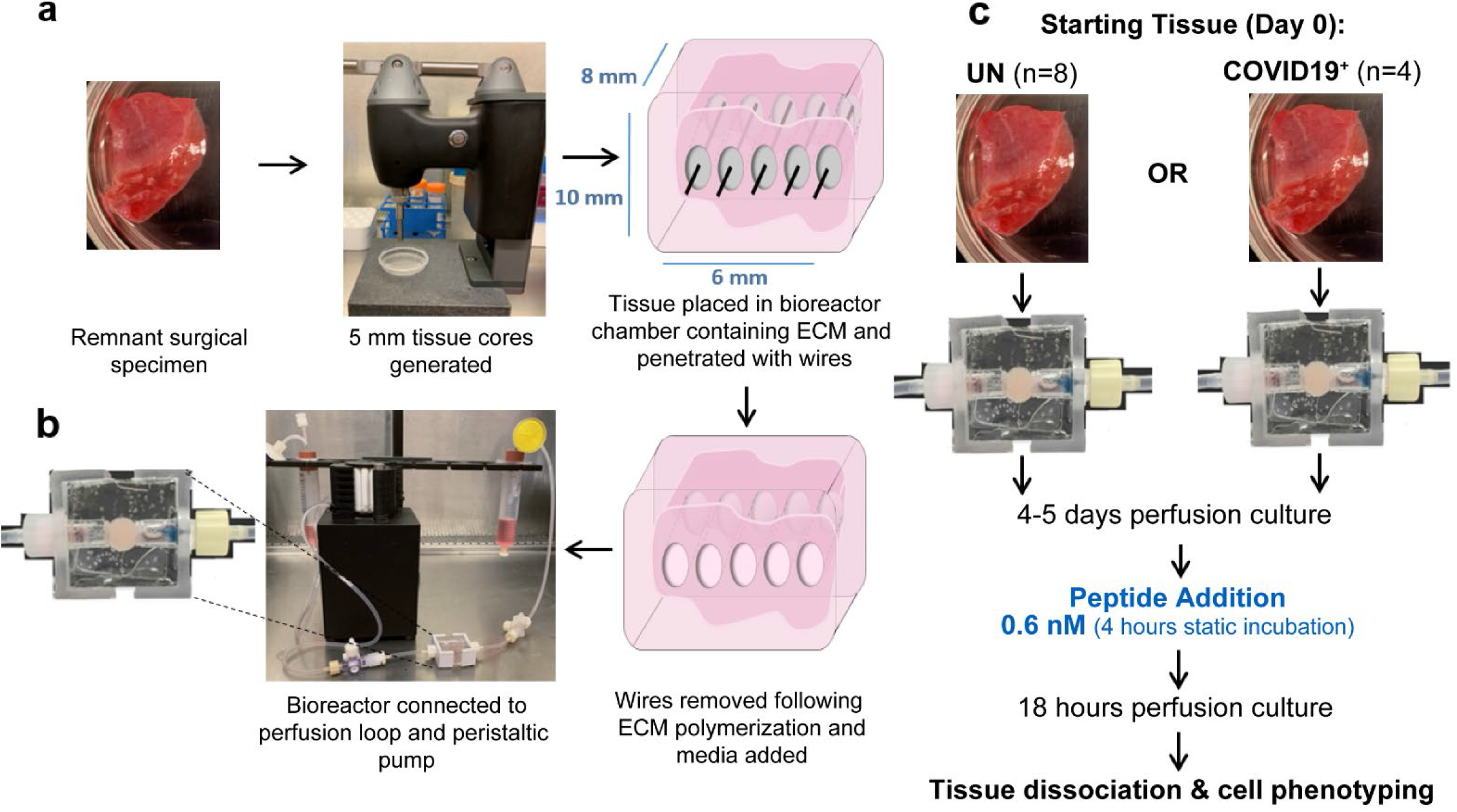
*Ex Vivo* Perfusion Culture of Human Lung and Exposure to SARS-CoV-2 Peptides. **A**. *Ex vivo* model setup process. **B**. Bioreactor chamber showing ECM volume containing tissue. **C**. Peptide exposure protocol.

### Cellular Landscape and Response to SARS-CoV-2 Peptides within the Lung Tissues of Previously Uninfected and Convalescent Individuals

Local immune responses to SARS-CoV-2 peptides were compared in lung tissue specimen collected from eight individuals who were not previously infected with SARS-CoV-2 (uninfected, UN) and four convalescent individuals who previously tested positive for SARS-CoV-2 and cleared the virus (COVID-19^+^), **Extended Data Tables 1 and 2** with subject demographics). Tissue cores were cultured *ex vivo* using the bioreactor platform and following four to five days perfusion culture, peptide pools covering the SARS-CoV-2 membrane glycoprotein (M peptide), nucleocapsid phosphoprotein (N peptide), or the immunodominant sequence of the spike protein (S peptide) were added to circulating media and cellular response was compared to vehicle control exposed tissues (**Figure 1C**).

While baseline phenotyping showed no difference in epithelial cell populations, endothelial cells, CD45^+^ immune cells, TNF-α^+^ immune cells, or CD4^+^ T cells within the lung tissues of COVID-19^+^ and UN individuals (**Extended Figs. 2A-C**, Extended Figs. 3A-B, Extended Fig. 4A, Figure 2A-B), a significant increase in % PD-1^+^ CD4^+^ T cells was noted (**Figure 2C**). Other subpopulations of CD4^+^ T cells showed no difference at baseline within the lungs of COVID-19^+^ and UN individuals (**Figure 2D, Extended Figs. 5A-E**). The frequency of CD8^+^ T cells was moderately increased in COVID-19^+^ compared to UN lung tissues (**Figure 2E**). Similar to CD4^+^ T cells, the PD-1^+^ CD8^+^ T cells were elevated in COVID-19^+^ at baseline (**Figure 2F**), but no differences were observed in other subpopulations of CD8^+^ T cells (**Figure 2G, Extended Figs. 6A-E**). The maintenance of cell populations over the culture period was assessed, comparing UN or COVID-19^+^ tissues in culture to the respective starting tissues. While the overall CD45^+^ frequencies declined over the culture period, the change in frequencies of epithelial, endothelial, T or B cell populations comparing UN and COVID-19^+^ lung tissues were not significantly different (**Extended Figs. 2D, 3C, 4B, 7, 8L-Y, & 9K-Q**); naïve CD8^+^ T cells showed a trending difference (p=0.068) following *ex vivo* culture (**Extended Fig 8S**).

**Figure 2:**
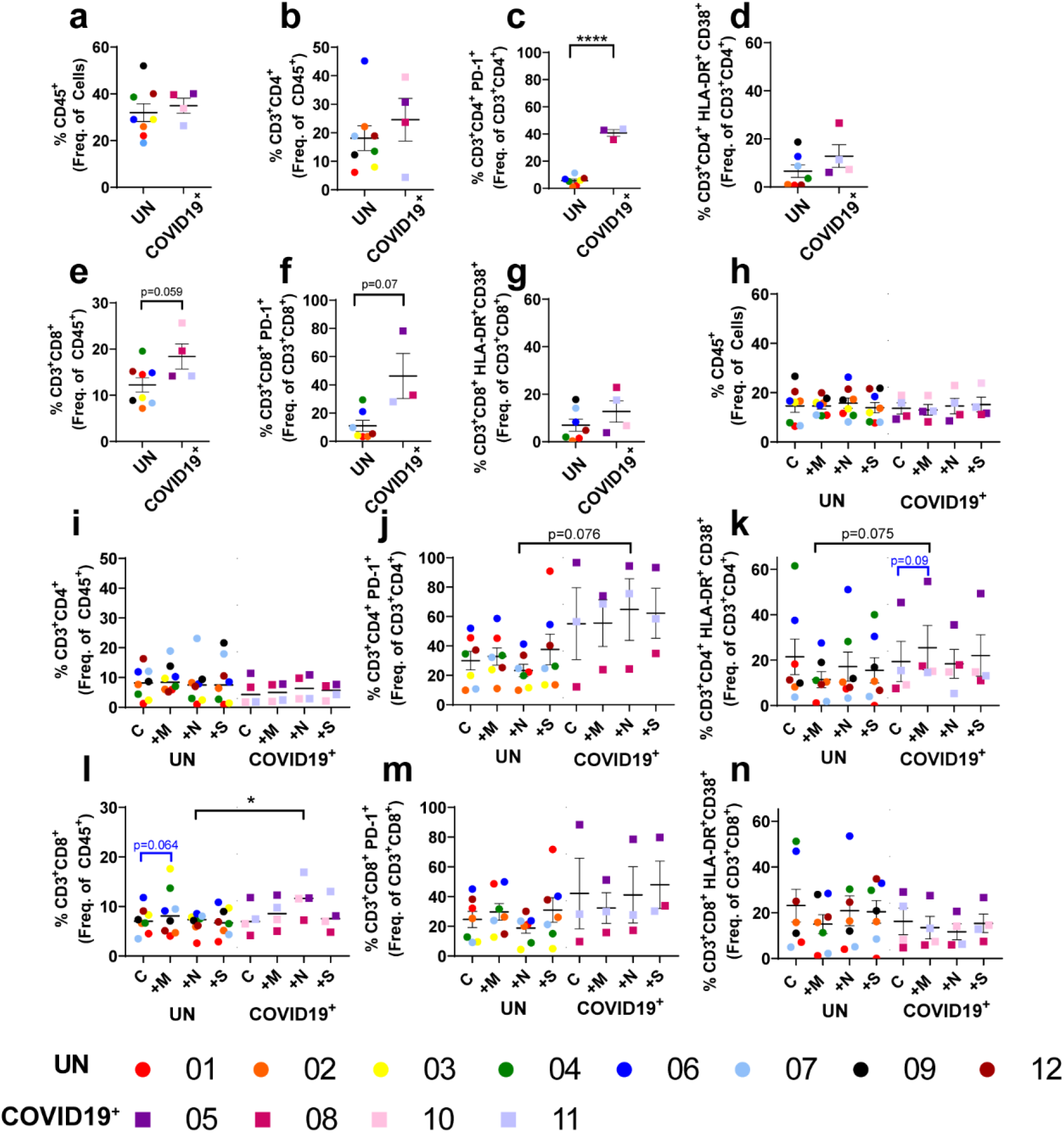
Local T Cell Response to SARS-CoV-2 Peptides within Lung Tissue Cores. **A-G**. Baseline T cell differences within lung tissue from uninfected individuals (UN) and individuals recovering from COVID-19 (COVID-19^+^). **H-N**. Impact of SARS-CoV-2 peptide exposure on T cell populations in UN and COVID-19^+^ lung tissue. n=8 UN and n=4 COVID-19^+^. Statistics shown in blue are comparisons between control and peptide exposed samples within each group (UN and COVID-19^+^). Statistics shown in black are the change in response between UN and COVID-19^+^ for each peptide when compared to the corresponding control.

Following exposure to the SARS-CoV-2 peptide pools, the lung tissue cores were collagenase digested and alterations in the cellular landscape and immune response were evaluated. As noted at baseline, no differences were observed in the epithelial cell populations, but an increase in CD31^+^ endothelial cells was found with M peptide treatment in COVID-19^+^ samples (**Extended Fig. 2E & Extended Fig. 3D**). No changes were noted in the CD45^+^ immune cells, TNF-α^+^ immune cells, or the CD4^+^ T cells with peptide exposure within UN or COVID-19^+^ lung tissues (**Figures 2H-I, Extended Fig. 4C**). The change in frequency of the PD-1^+^ CD4^+^ T cells between control and N peptide-exposed tissues was greater in COVID-19^+^ compared to UN lung tissues (**Figure 2J**). Additionally, hyperactivated CD4^+^ T cells showed an increasing trend in COVID-19^+^ tissues (p=0.075) exposed to the M peptide when compared to control; this change was not observed in UN tissues (**Figure 2K**). No significant changes were noted in proliferating, antigen specific, or IFN-γ^+^CD4^+^ T cells (**Extended Figs. 5F-I**). The frequency of CD8^+^ T cells increased in response to the M peptide (p=0.064) in UN tissues when compared to control. This population also showed a significant increase when N peptide-response was compared between UN and COVID-19^+^, with increased CD8^+^ T cells in COVID-19^+^ (**Figure 2L**). The frequency of proliferating and antigen specific CD8^+^ T cells, although not significant (*p* = 0.051 with M peptide exposure and *p* = 0.093 with S peptide exposure, respectively), increased with peptide exposure in UN, but not in the COVID-19^+^ tissues (**Extended Figs. 6F-G**). No significant differences were observed in PD-1^+^, hyperactivated, or IFN-γ^+^CD8^+^ T cells (**Figures 2M-N, Extended Figs. 6H-I**).

### Memory T cell Response to SARS-CoV-2 Peptides within the Lung Tissues of Convalescent Individuals

We assessed T cell memory subsets within the lung tissues of UN and COVID-19^+^ individuals, as they play a vital role in viral clearance during re-infection and recent studies identified functional memory T cells within the peripheral blood of COVID-19^+^ patients^22,24-29^. At baseline, CD4^+^ or CD8^+^ lung memory T cell subsets were not significantly different comparing UN and COVID-19^+^ tissues (**Figure 3A-J, Extended Figs. 8A-G**). However, tissue resident memory (TRM) and effector memory (EM) were the most prevalent subsets within the lung tissue (**Figures 3A, 3F, 3E & 3J)**. Following *ex vivo* culture, TRM responses to all peptide pools were noted within the UN tissues, with decreasing CD4^+^ TRM frequency with peptide exposure (**Figure 3K**). We determined the % IFN-γ^+^ subsets within the CD4^+^ and CD8^+^ TRM as well as % TRM within the overall IFN-γ^+^ CD4^+^ and CD8^+^ cells. Although statistical significance was not observed when comparing UN and COVID-19^+^, patient-specific differences were noted in COVID-19^+^. At baseline, the average % IFN-γ^+^ CD8^+^TRM in COVID-19^+^ was 1.71 fold higher than UN average (**Figure 3G)**. Within the COVID-19^+^, % IFN-γ^+^ CD8^+^TRM and % IFN-γ^+^ CD4^+^TRM in Subject #10 who received COVID-19 vaccination, were 3.69 and 4.65 fold higher, respectively, than average of UN (**Figure 3Q, 3L)**. While meaningful fold change in average %TRM within CD8^+^IFN-γ^+^ was not noted in this individual, the average %TRM within CD4^+^IFN-γ^+^ in COVID-19^+^ was 2 fold higher than UN average (**Figure 3M)**. Of the total IFN-γ^+^ CD8^+^ T cells in the lungs of COVID-19^+^ individuals #11 (COVID^+^ twice) and #8, 40-50% were TRM (**Figure 3R**). Both % IFN-γ^+^ of the CD8^+^TRM (2.55-fold) and % TRM within CD4^+^IFN-γ^+^ (2.26-fold) were higher in Subject #11 (**Figure 3R, 3L**). In COVID-19^+^ Subject #10, the % IFN-γ^+^ CD8^+^TRM was 1.5 fold higher in response to the S peptide when compared to control (**Figure 3Q**), and the %TRM within CD8^+^IFN-γ^+^ cells was 3.25 fold and 2.72 fold higher in M-peptide and N-peptide exposed tissues, respectively (**Figure 3R**). Interestingly, in COVID-19^+^ #5, % IFN-γ^+^ CD8^+^TRM was 5.88 fold and 10 fold higher in the S-peptide and M-peptide treated samples compared to the respective controls; robust response was not noted with N-peptide (**Figure 3Q**). The % TRM CD4^+^IFN-γ^+^ in COVID-19^+^ #5 was 12.5 fold higher in response to the S peptide when compared to control, whereas other COVID-19^+^ did not demonstrate a robust response in (**Figure 3M**); in COVID-19^+^ #8, the M-peptide response was 3.39 fold higher compared to control (**Figure 3M**). The % IFN-γ^+^ within the CD4^+^TRM cells that respond to the S peptide were 14.3 fold higher compared to control in COVID-19^+^ #5, while this response was minimal in COVID-19^+^ #10 and 11 (**Figure 3L**). But the % IFN-γ producing CD4^+^TRM were 2.87 fold higher in response to the M peptide and were 3.31 fold higher in response to the N peptide in vaccinated COVID-19^+^ #10 and COVID-19^+^ #11 (COVID^+^ twice) respectively, compared to control (**Figure 3L**).

**Figure 3:**
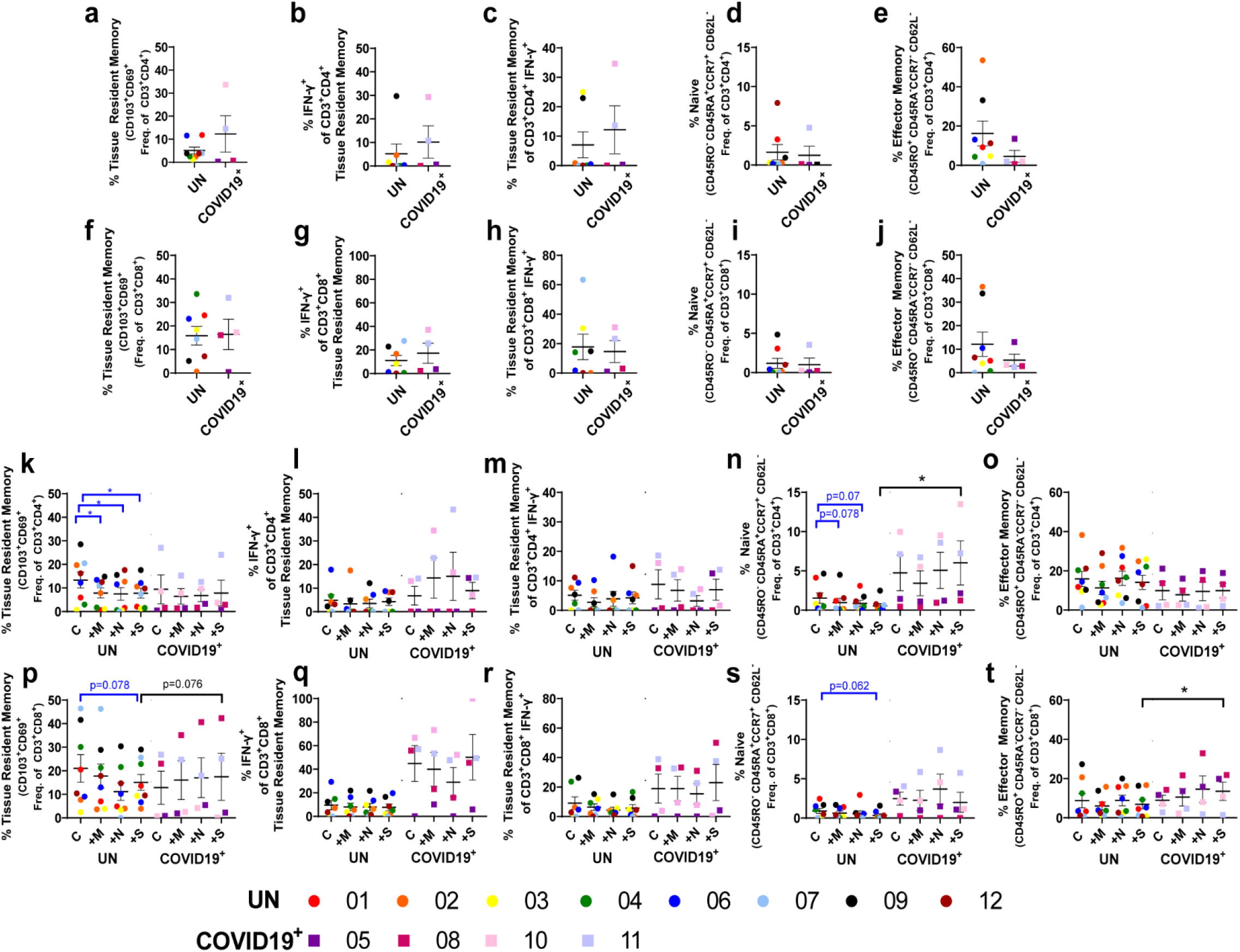
Memory T Cell Response to SARS-CoV-2 Peptides within the Lung. **A-J**. Baseline differences in memory T cells within lung tissue from uninfected individuals (UN) and individuals recovering from COVID-19 (COVID-19^+^). **K-T**. Impact of SARS-CoV-2 peptide exposure on memory T cell populations in UN and COVID-19^+^ lung tissue. n=8 UN and n=4 COVID-19^+^. Statistics shown in blue are comparisons between control and peptide exposed samples within each group (UN and COVID-19^+^). Statistics shown in black are the change in response between UN and COVID-19^+^ for each peptide when compared to the corresponding control.

Additionally, stem cell like CD4^+^ memory T cells increased (p=0.078) with M peptide exposure (**Extended Fig. 8H**), whereas both naïve and central memory CD4^+^ T cell frequencies decreased with M-peptide exposure; a similar reduction in N peptide response within naïve CD4^+^ T cells was noted in UN tissues (**Figures 3N, Extended Fig. 8I**). Interestingly, the change in the frequency of naïve CD4^+^ T cells, when comparing the control and S peptide response, was significantly increased in COVID-19^+^ when compared to UN (**Figure 3N**). No significant differences were observed in CD4^+^ EM cells (**Figure 3O**). Amongst CD8^+^ memory subsets, a trend towards a reduction in S peptide-responding TRM CD8^+^ T cells was noted (p=0.076), when compared to respective controls in UN tissues. This trend coincided with an increasing trend in the change in % TRM between control and S peptide exposure when comparing UN and COVID-19^+^; this cell population increased with peptide stimulation in COVID-19^+^ tissues (**Figure 3P**). No differences were observed in CD8^+^ stem cell like memory T cells and minimal changes were observed in CD8^+^ naïve and central memory subsets (**Extended Fig. 8J-K, Figure 3S**). Importantly, CD8^+^ EM cells tended to increase with peptide exposure in COVID-19^+^ tissues, with a significant increase in the change between control and S peptide treated samples in COVID-19^+^ compared to UN tissues (**Figure 3T**).

As the humoral immune response to infection is essential for protection and has not been evaluated in SARS-CoV-2 convalescence at the tissue level, we evaluated B cell subsets in a subset of the UN and COVID-19^+^ lung tissues. No significant differences in B cell subsets were noted at baseline (**Figure 4A-C, Extended Figs. 9A-F**). A trend towards an increase in CD19^+^ B cells following *ex vivo* culture and S peptide exposure (p=0.068) was observed in COVID-19^+^ tissues when compared to control. Furthermore, the change between control and S peptide exposed samples was significantly different in COVID-19^+^ when compared to UN tissues (**Figure 4D**). When evaluating memory B cells, opposing changes between control and M peptide exposed samples were noted in UN and COVID-19^+^ (**Extended Fig. 9H**). While transitional B cells were boosted in UN, plasmablasts tended to be higher in COVID-19^+^ samples; all other B cell subsets remained unchanged with peptide exposure (**Figures 4E-F, Extended Figs. 9G and 9I-J**). Evaluation of humoral responses in the lung tissue supernatants showed that IgA levels were higher at baseline in 2/4 recovering COVID-19^+^ (#11, infected twice and unvaccinated) and # 8 (infected once and unvaccinated, see Extended Table 2) compared to UN (**Figure 4G)**. The IgA response was more dominant than the IgG response in these lung tissue supernatants of COVID-19^+^ (**Figure 4H)**. Additionally, while the baseline IgA response was not detectable in COVID-19^+^ #5 (infected once and unvaccinated), both M and N peptide stimulation increased IgA levels substantially **(Figure 4G)**. Similarly, while baseline IgA was also low in COVID-19^+^ # 10 (vaccinated after first infection), both M and N peptide stimulation increased IgA levels. We then used confocal microscopy to identify CD138^+^ plasma cells secreting IgA in the lungs of COVID-19^+^ compared to UN. Consistent with SARS-CoV-2 specific IgA levels, we detected more CD138^+^IgA^+^ cells in the lungs of COVID-19^+^ individuals at baseline and in response to SARS-CoV-2-specific peptides **(Figures 4I and Extended Fig. 10)**.

**Figure 4:**
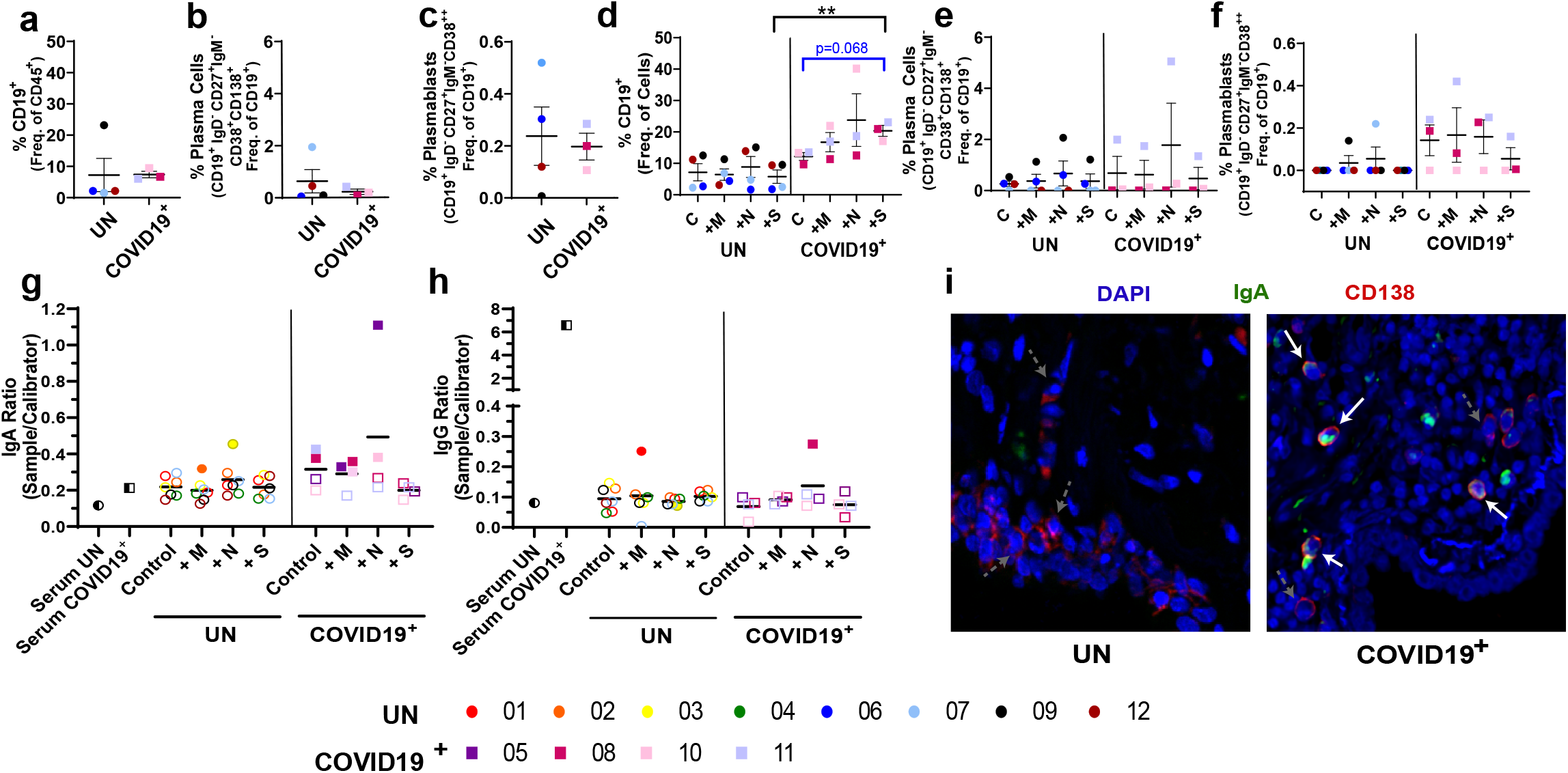
Local B Cell Response to SARS-CoV-2 Peptides within the Lung. **A-C**. Baseline differences in B cell populations within lung tissue from uninfected individuals (UN) and individuals recovering from COVID-19 (COVID-19^+^). **D-F**. Impact of SARS-CoV-2 peptide exposure on B cell populations in UN and COVID-19^+^ lung tissue. **G-H**. Quantification of circulating SARS-CoV-2 specific IgA (**G**) and IgG (**H**), opaque symbols indicate positive samples (measurements over threshold). **I**. Photomicrographs showing CD138^+^ plasma cells (red) secreting IgA (green) in UN (left) and COVID-19^+^ samples (right) at day 0 (starting tissue). White arrows pointing to CD138^+^ plasma cells secreting IgA, grey dashed arrows pointing to CD138^+^plasma cells without IgA secretion. n=4 UN and n=3 COVID-19^+^. Statistics shown in blue are comparisons between control and peptide stimulated samples within each group (UN and COVID-19^+^). Statistics shown in black are the change in response between UN and COVID-19^+^ for each peptide when compared to the corresponding controls.

## CONCLUSIONS

T cell dynamics during both the acute and memory phases following SARS-CoV-2 infection determine the specific T cell and humoral responses optimized for host protection. Specifically, the role of SARS-CoV-2 specific effector T cells on viral clearance and their accumulation in COVID-19^+^ and recovered individuals over time requires further study; however, modeling local immune responses against SARS-CoV-2 has posed challenges. We utilized a novel three-dimensional perfused human lung tissue model that maintains cellular heterogeneity, viability and extracellular matrix components over an extended culture period. In this model, we evaluated local immune responses to SARS-CoV-2 peptide pools that represent membrane, nucleocapsid and spike proteins of SARS-CoV-2. We provide evidence for pre-existing T cell immunity and SARS-CoV-2 peptide-specific local T and B lymphocyte memory responses in lung tissues from COVID-19^+^ and uninfected (UN) individuals.

Evidence of elevated frequencies of PD-1^+^ CD4^+^ and/or CD8^+^ T cells in our COVID-19^+^ lung tissue models are consistent with the reported increase of circulating PD-1^+^ cells in COVID-19^+^ patients ^40-43^, suggesting that these models are relevant for evaluation of antigen primed responses. Pre-existing T cell immunity in UN lungs, however, was represented by M-peptide response in CD8^+^ stem cell like memory cells, and not N-peptide responding CD8^+^ T cells as reported earlier in circulation ^6,7,14-23^. Interestingly, the change in frequency of overall N-peptide responding CD8^+^ T cells was increased significantly in COVID-19^+^ compared to UN. Additionally, the change in N-peptide responding PD-1^+^ CD4^+^T cells was significantly higher in COVID-19^+^ compared to UN. While these changes are not consistent with what is reported in circulation, the differences in the lung niche may impact local immune responses differently from what is seen in circulation.

Previous studies in SARS-CoV recovered individuals have identified persistent memory T cells that suggested vaccine-mediated induction of TRMs may provide long-term protection against this pandemic^12^. Consistent with this, a larger proportion of CD8^+^T cell EM cells and those with TRM phenotype have been identified in bronchoalveolar lavage fluid and lung tissues of individuals with COVID-19^44,45^. A key role for TRMs in protection against pathogen challenge has been established for many tissues, including the lung^12,44-46^. We report here, the existence and long-term maintenance in culture of a high frequency of SARS-CoV-2-S-peptide specific EM CD8^+^ T cells and TRM in COVID-19^+^ lungs. In our studies, 3 out of the 4 COVID-19^+^ showed robust increase in % IFN-γ^+^TRM. While the vaccinated COVID-19^+^ did not demonstrate a significant increase compared to other COVID-19^+^ at baseline, an IFN-γ^+^ S-peptide response in TRMs was noted in this tissue. Further, fold change in % TRM of the total IFN-γ^+^ cells which showed M and N-peptide responses were increased in the vaccinated COVID-19^+^. These observations are consistent with vaccine mediated induction of TRMs as a potential long term protection strategy^12,47^. Additionally, vaccine induced TRMs localized to lung tissue of COVID-19^+^ suggest potential beneficial effects in the respiratory tract. Further, IFN-γ secretion by CD8^+^ T cells was predominantly in response to S-peptide in COVID-19^+^ and the overall TRM response to S-peptide was higher in COVID-19^+^. In this context, high frequencies of spike protein-specific CD4^+^T cell responses have been reported in blood of COVID-19 convalescent ^7,14,15,28,48^. Importantly, CD4^+^T cells are necessary for the formation of protective CD8^+^TRM during influenza infection; IFN-γ is an essential signal for this process^49^. Consistent with this, despite the small size in our study, the %TRM within CD4^+^ IFN-γ producers was increased in COVID-19^+^ ; a robust 12.5 fold increase in S-peptide responder TRMs was identified within CD4^+^ cells in one COVID-19^+^. Additionally, both IFN-γ^+^CD4^+^ and CD8^+^ TRM are responding to both M and N-peptides in addition to S-peptide pools. These data suggest that inclusion of nucleocapsid as well as other structural viral proteins in vaccine approaches may broaden and balance the functional profile of memory T cells, resembling control of natural infection.

Additionally, we report for the first time, B cell responses to SARS-CoV-2 peptide pools against spike protein in COVID-19^+^ lung tissues, with a significant change in % S-peptide responding CD19^+^ cells in COVID-19^+^ compared to UN lung tissues; it remains to be determined if the S-peptide responders are tetramer^+^ antigen-reactive B cells. The increased immature B cell frequencies in the COVID-19^+^ lungs may represent what is in circulation, as lung tissues were not perfused prior to *ex vivo* culture. Importantly, we report increased IgA levels and IgA^+^ plasma cells in the lungs of COVID-19^+^ compared to UN, either at baseline or in response to SARS-CoV-2 peptides. The small sample size may prevent us from discerning associations between vaccination or infection status and/or IgA presence. As secretory IgA plays a crucial role in protecting mucosal surfaces against respiratory viruses and other pathogens, vaccine approaches either by nebulization or intranasal delivery may enable enhancement of IgA in the upper airways and afford long term protection against COVID-19.

## METHODS

### Clinical Sample Collection

De-identified, remnant surgical specimen were obtained from lobectomy and wedge resection surgeries performed at the University of Alabama at Birmingham. For peptide exposure studies, 12 tissue specimen were obtained from patients with no history of SARS-CoV-2 infection and 4 tissue specimen were obtained from patients who had previously tested positive for SARS-CoV-2 and cleared the infection. This study was approved by the University of Alabama at Birmingham Institutional Review Board (IRB-300003092 and IRB-300003384) and conducted following approved guidelines and regulations. Written informed consent was obtained from all participants. Patient demographics are described in **Table 1**.

### Sample Processing and *Ex Vivo* Perfusion Culture

5 mm diameter tissue cores were generated from remnant surgical specimen using a tissue coring press (Alabama Research and Development, USA). One tissue core was placed into the central chamber of a polydimethylsiloxane (PDMS, Krayden, USA) bioreactor containing a mixture of extracellular matrix (ECM, 90% collagen type 1 (Advanced Biomatrix, USA) + 10% growth factor reduced Matrigel (Corning, USA)) components for structural support as previously described^50^. The tissue/ECM volume was then penetrated with five 400 micron Teflon coated stainless steel wires to generate through-channels for tissue perfusion. Following ECM polymerization, the wires were removed and the through-channels were filled with tissue culture media (1:1 mixture of X-Vivo15 and Bronchial Epithelial Growth media (Lonza, USA) with antibiotics (MP Biomedicals, USA)). The bioreactor was then connected to a perfusion system, that contained a media reservoir, peroxide cured silicon tubing (Cole Parmer, USA), a collection reservoir and peristaltic pump (ESI, USA), and tissue culture media was perfused through the tissue volume for 5 to 14 days (37°C, 5% CO_2_), with media changed every 3 days. At the end of each experiment, a portion of each tissue was fixed separately for histologic processing and collagenase B (Roche, Switzerland) digestion for flow cytometry analysis

### SARS-CoV-2 Peptide Exposures

For *in vitro* peptide exposures, on day 5 of culture, conditioned media was collected from the collection reservoir and 0.6 nM SARS-CoV-2 peptides (Peptivator peptide pools: M, N and S, Miltenyi Biotec, USA) or vehicle control (cell culture grade water) were added to the tissue chamber of the bioreactor per manufacturer instructions. Following a four hour static incubation, tissue culture media containing brefeldin A and monensin (BD, Germany) was perfused throughout the tissue chamber for an additional 14 hours.

### Multiparametric Flow Cytometry

#### The following antibodies were used for multiparametric flow cytometry for T cell analysis

Anti-CD3-alexafluor 700 (Clone: UCHT1); anti-CD4-FITC (Clone: RPA-T4); anti-CD69-BV563 (Clone: FN50) from BD Biosciences (Germany). Anti-HLA-DR-APC (Clone: LN3); anti-CD3-PE-Cy7 (Clone: UCHT1); anti-CD8-APC (Clone: 53-6.7); anti-Interferon-γ-Alexafluor700 (Clone:B27) from eBioscience (Thermo Fisher, Germany). Anti-Ki-67-Dylight350 (Clone: 1297A) from Novus (USA). Anti-CD45-APC-Cy7 (Clone:2D1); anti-CCR7-Pacific Blue (Clone: G043H7); anti-CD45RA-BV510 (Clone:HI100); anti-CD45RO-PerCy-Cy5.5 (Clone:UCHL1); anti-CD62L-BV650 (Clone:DREG-56); anti-CD103-PE (Clone:Ber-ACT8): anti-CD8-BV510 (Clone:SK1); anti-CD38-PE-Cy7 (Clone:HB-7); anti-CD154-PE/Dazzle (Clone:24-31) anti-PD-1-BV605 (Clone:NAT105); anti-TNF-αBV605 (Clone:MAb11) from Biolegend (USA).

#### The following antibodies were used for multiparametric flow cytometry for analysis of resident immune and structural cells

Anti-CD64-PerCp-eFluor710 (Clone: 10.1); anti-CD11b-APC-Cy7 (Clone: ICRF44); anti-HLA-DR-FITC (Clone: LN3); anti-EpCAM(CD326)-Alexafluor 594 (Clone:9C4) from eBioscience (Thermo Fisher, Germany). Anti-CD16-PE (Clone: 3G8); anti-CD45-Pacific Blue (Clone: HI30); anti-CD66b-PerCP-Cy5.5 (Clone: G10F5) from Biolegend (USA). Anti-CD31-Alexafluor 700 (Clone: UCHT1) from BD Biosciences (Germany) anti-PanCytokerain-APC (Clone: C-11) and anti-Ki-67-Dylight350 (Clone: 1297A) from Novus Biologicals (USA).

#### The following antibodies were used for multiparametric flow cytometry for B cell analysis

anti-CD10-BV650 (Clone:HI10A); anti-CD19-eFluor450 (Clone:HIB19) from eBioscience (Thermo Fisher, Germany). Anti-CD45-BV605 (Clone: 2D1); anti-CD21-PerCP-Cy5 (Clone:Bu32); anti-CD24-APC-Cy7 (Clone:ML5); anti-CD27-APC (Clone: M-T271); anti-CD38-PE-Cy7 (Clone: HB-7); anti-CD138-BV510 (Clone:MI15); anti-IgD-FITC (Clone:IA6-2); anti-IgM-PE (Clone:MHM-88) from Biolegend (USA). The Foxp3 / Transcription Factor Staining Buffer Set (Thermo Fisher, Germany) and the Cytofix/Cytoperm Fixation/Permeablization kit (BD, Germany) were used according to the manufacturer’s protocol to stain for intracellular molecules (intranuclear and cytoplasmic molecules, respectively). Analyses were performed on FACSymphony A3 Cell Analyzer with FACSDiva software version 8.0.1 (BD Biosciences, Germany). Data were analyzed with FlowJo 10.7.1 (Treestar, USA).

### Analysis of IgA and IgG

Analysis of secreted Sars-CoV-2 specific IgA and IgG was performed using conditioned media and serum along with ELISA kits (EuroImmun). Conditioned media was concentrated using Amicon Ultra-4 100 K Centrifugal Filter Units (Millipore Sigma), prior to analysis. Samples were spun at 4000 x g for 15 minutes using a swinging bucket centrifuge, per manufacture’s recommendation. ELISA assays were performed following the manufacturer’s recommendations (1:101 dilution of serum samples and 1:5 dilution of conditioned media samples) and a threshold was set to determine positive levels of immunoglobulin based on the average level of immunoglobulin in control media samples.

### Histologic Processing and Analysis

Following *ex vivo* culture, a portion of each cultured tissue was fixed with neutral buffered formalin, processed to paraffin, and histological (FFPE) sections were prepared, as previously described^51^. 5 micron sections were stained with hematoxylin and eosin (H&E) to evaluate tissue morphology and cell density (number of cells per cross-sectional area) as described before^52^.

### Immunofluorescence Staining and Analysis

Immunofluorescence staining was performed on FFPE sections to detect plasma cells secreting IgA. Anti-CD138 (Syndecan-1, 1:50, clone aa18-218, LSBio) was detected with anti-mouse Alexafluor 647 (1:500 Life Technologies) and IgA (1:100, clone Mc24-2E11, LSBio) was detected with anti-rabbit Alexafluor-594 (1:500, Life Technologies), following antigen retrieval (10□mM citrate buffer, pH 6). 4′,6-Diamidino-2-phenylindole (DAPI) (1:1000) was used as a nuclear counterstain. Sections were imaged using a Nikon A1R-HD25 confocal microscope with a Plan Apo λ 10x objective (na .5 wd 4000).

### Matrix Proteomics

For matrix protein enrichment and extraction, tissue samples were prepared as described before^53^. Briefly, tissues were processed using the Millipore Compartment Protein Extraction Kit with some modifications of the described methodology^53^ and all fractions were stored at -80° overnight. The ECM fraction was then reconstituted in 8M urea and deglycosylated. The urea-insoluble fraction was collected by centrifugation, reconstituted in 1x LDS sample buffer, and sonicated for 20 minutes in an ultrasonic water bath. Both urea-soluble and insoluble fractions were quantified via EZQ protein assay, and an equal amount per sample was loaded onto 10% Bis-tris gels and gels were stained overnight with Colloidal Coomassie. Each sample was then digested in 3 fractions with trypsin overnight and high resolution LC-ESI-MS/MS analysis was completed. Data was searched against human subset of Uniref100 database with Carbamidomethylation, Oxidation, and Hydroxyproline.

### Measuring Lactate Dehydrogenase

Lactate dehydrogenase (LDH) was measured in conditioned media using the Invitrogen CyQUANT LDH Cytotoxicity Assay (Thermo Fisher, Germany) following manufacturer’s instruction.

### Statistical Analysis

The measured flow data was summarized by presenting descriptive statistics, such as mean with standard error of the mean (SEM), in the uninfected and convalescent groups. Changes of the measurement between control and each of peptides were computed separately. Two sample t-tests and Wilcoxon rank-sum tests were performed to determine if means of the changes were different between uninfected and convalescent groups. Mean and SD of the outcome measured were estimated by control and peptides within uninfected and convalescent groups respectively. To evaluate difference in the outcome between control and each of peptides within each of the groups, paired t-test and Wilcoxon signed-rank tests were used. All other statistical analyses were performed using SAS 9.4 (SAS Institute, USA). Statistical significance was determined at P-value < 0.05.

## Data Availability

The authors confirm that the data supporting the findings of this study are available within the article [and/or] its supplementary materials.

## ACKNOWLEDGEMENTS

Research reported in this publication was supported by the University of Alabama at Birmingham School of Medicine COVID-19 pilot grant and P42 ES027723 (Project 2) awarded to J.S.D., UAB Comprehensive Flow Cytometry Core Facility (NIH P30 AR048311 and NIH P30 AI27667), the UAB Tissue Biorepository, the UAB Pathology Core Research Laboratories and the UAB High Resolution Imaging Facility. The authors would like to thank Dr. Paul Goepfert for providing insights in T cell immunology in infection and Dr. Maite Sabalza for expertise in IgA/IgG ELISA.

## AUTHOR CONTRIBUTIONS

K.F.G. was involved in bioreactor model set up, experimental design and execution, data collection, data analysis, and manuscript and figure preparation. C.S.S., S.A.K., and A.M.W. were involved in data collection, data analysis, and manuscript editing. Y.W. was involved in data collection and manuscript editing. J.L.B. was involved in bioreactor design and production and manuscript editing. J.A.M. was involved in the design and analysis of ECM proteomics and manuscript editing. Y.K. was completed statistical analyses and was involved in manuscript editing. V.J.T. and K.S.H were involved in experimental design and reviewing the overall concepts and thought process of the manuscript. M.A. gave critical insights for the manuscript. J.M.D. facilitated collection of remnant surgical specimen and was involved in manuscript editing. J.S.D. was involved in experimental design and oversight, data interpretation, and manuscript and figure preparation.

Correspondence and requests for materials should be addressed to Jessy Deshane

## REFERENCES

1 BioSpace. Enzo Announces Issuance of U.S. Patent for Methods of Using Proprietary Compound SK1-I in Patients; Exploring Options for Development as a Potential Treatment for COVID-19. (2020). <https://www.biospace.com/article/releases/enzo-announces-issuance-of-u-s-patent-for-methods-of-using-proprietary-compound-sk1-i-in-patients-exploring-options-for-development-as-a-potential-treatment-for-covid-19/>.

2 Control, C. F. D. United States COVID-19 Cases, Deaths, and Laboratory Testing (NAATs) by State, Territory, and Jurisdiction. <https://www.cdc.gov/coronavirus/2019-ncov/cases-updates/cases-in-us.html.>.

3 Worldometer. COVID-19 Coronavirus Pandemic (2022). <https://www.worldometers.info/coronavirus/#countries>.

4 De Biasi, S. et al. Marked T cell activation, senescence, exhaustion and skewing towards TH17 in patients with COVID-19 pneumonia. Nat Commun 11, 3434, doi:10.1038/s41467-020-17292-4 (2020).

5 de Candia, P., Prattichizzo, F., Garavelli, S. & Matarese, G. T Cells: Warriors of SARS-CoV-2 Infection. Trends Immunol 42, 18–30, doi:10.1016/j.it.2020.11.002 (2021).

6 Files, J. K. et al. Sustained cellular immune dysregulation in individuals recovering from SARS-CoV-2 infection. J Clin Invest 131, doi:10.1172/JCI140491 (2021).

7 Le Bert, N. et al. SARS-CoV-2-specific T cell immunity in cases of COVID-19 and SARS, and uninfected controls. Nature 584, 457–462, doi:10.1038/s41586-020-2550-z (2020).

8 Toor, S. M., Saleh, R., Sasidharan Nair, V., Taha, R. Z. & Elkord, E. T-cell responses and therapies against SARS-CoV-2 infection. Immunology 162, 30–43, doi:10.1111/imm.13262 (2021).

9 Farinholt, T. et al. Transmission event of SARS-CoV-2 Delta variant reveals multiple vaccine breakthrough infections. medRxiv, doi:10.1101/2021.06.28.21258780 (2021).

10 Olsen, R. J. et al. Trajectory of Growth of SARS-CoV-2 Variants in Houston, Texas, January through May 2021 Based on 12,476 Genome Sequences. Am J Pathol, doi:10.1016/j.ajpath.2021.07.002 (2021).

11 Zhao, J. et al. Airway Memory CD4(+) T Cells Mediate Protective Immunity against Emerging Respiratory Coronaviruses. Immunity 44, 1379–1391, doi:10.1016/j.immuni.2016.05.006 (2016).

12 Channappanavar, R., Fett, C., Zhao, J., Meyerholz, D. K. & Perlman, S. Virus-specific memory CD8 T cells provide substantial protection from lethal severe acute respiratory syndrome coronavirus infection. J Virol 88, 11034–11044, doi:10.1128/JVI.01505-14 (2014).

13 Zhao, J., Zhao, J. & Perlman, S. T cell responses are required for protection from clinical disease and for virus clearance in severe acute respiratory syndrome coronavirus-infected mice. J Virol 84, 9318–9325, doi:10.1128/JVI.01049-10 (2010).

14 Grifoni, A. et al. Candidate Targets for Immune Responses to 2019-Novel Coronavirus (nCoV): Sequence Homology-and Bioinformatic-Based Predictions. SSRN, 3541361, doi:10.2139/ssrn.3541361 (2020).

15 Grifoni, A. et al. Targets of T Cell Responses to SARS-CoV-2 Coronavirus in Humans with COVID-19 Disease and Unexposed Individuals. Cell 181, 1489–1501 e1415, doi:10.1016/j.cell.2020.05.015 (2020).

16 Nelde, A. et al. SARS-CoV-2-derived peptides define heterologous and COVID-19-induced T cell recognition. Nat Immunol 22, 74–85, doi:10.1038/s41590-020-00808-x (2021).

17 Schulien, I. et al. Characterization of pre-existing and induced SARS-CoV-2-specific CD8(+) T cells. Nat Med 27, 78–85, doi:10.1038/s41591-020-01143-2 (2021).

18 Kared, H. et al. CD8+ T cell responses in convalescent COVID-19 individuals target epitopes from the entire SARS-CoV-2 proteome and show kinetics of early differentiation. bioRxiv, doi:10.1101/2020.10.08.330688 (2020).

19 Meckiff, B. J. et al. Imbalance of Regulatory and Cytotoxic SARS-CoV-2-Reactive CD4(+) T Cells in COVID-19. Cell 183, 1340–1353 e1316, doi:10.1016/j.cell.2020.10.001 (2020).

20 Ong, E. Z. et al. A Dynamic Immune Response Shapes COVID-19 Progression. Cell Host Microbe 27, 879–882 e872, doi:10.1016/j.chom.2020.03.021 (2020).

21 Parveen, F. et al. Role of Ceramidases in Sphingolipid Metabolism and Human Diseases. Cells 8, doi:10.3390/cells8121573 (2019).

22 Rydyznski Moderbacher, C. et al. Antigen-Specific Adaptive Immunity to SARS-CoV-2 in Acute COVID-19 and Associations with Age and Disease Severity. Cell 183, 996–1012 e1019, doi:10.1016/j.cell.2020.09.038 (2020).

23 Sattler, A. et al. SARS-CoV-2-specific T cell responses and correlations with COVID-19 patient predisposition. J Clin Invest 130, 6477–6489, doi:10.1172/JCI140965 (2020).

24 Peng, Y. et al. Broad and strong memory CD4(+) and CD8(+) T cells induced by SARS-CoV-2 in UK convalescent individuals following COVID-19. Nat Immunol 21, 1336–1345, doi:10.1038/s41590-020-0782-6 (2020).

25 Rodda, L. B. et al. Functional SARS-CoV-2-Specific Immune Memory Persists after Mild COVID-19. Cell 184, 169–183 e117, doi:10.1016/j.cell.2020.11.029 (2021).

26 Jung, J. H. et al. SARS-CoV-2-specific T cell memory is sustained in COVID-19 convalescent patients for 10 months with successful development of stem cell-like memory T cells. Nat Commun 12, 4043, doi:10.1038/s41467-021-24377-1 (2021).

27 Lipsitch, M., Grad, Y. H., Sette, A. & Crotty, S. Cross-reactive memory T cells and herd immunity to SARS-CoV-2. Nat Rev Immunol 20, 709–713, doi:10.1038/s41577-020-00460-4 (2020).

28 Ni, L. et al. Detection of SARS-CoV-2-Specific Humoral and Cellular Immunity in COVID-19 Convalescent Individuals. Immunity 52, 971–977 e973, doi:10.1016/j.immuni.2020.04.023 (2020).

29 Tavukcuoglu, E., Horzum, U., Cagkan Inkaya, A., Unal, S. & Esendagli, G. Functional responsiveness of memory T cells from COVID-19 patients. Cell Immunol 365, 104363, doi:10.1016/j.cellimm.2021.104363 (2021).

30 Shaan Lakshmanappa, Y. et al. SARS-CoV-2 induces robust germinal center CD4 T follicular helper cell responses in rhesus macaques. Nat Commun 12, 541, doi:10.1038/s41467-020-20642-x (2021).

31 Grigoryan, L. & Pulendran, B. The immunology of SARS-CoV-2 infections and vaccines. Semin Immunol 50, 101422, doi:10.1016/j.smim.2020.101422 (2020).

32 Sette, A. & Crotty, S. Pre-existing immunity to SARS-CoV-2: the knowns and unknowns. Nat Rev Immunol 20, 457–458, doi:10.1038/s41577-020-0389-z (2020).

33 Gaebler, C. et al. Evolution of antibody immunity to SARS-CoV-2. Nature 591, 639–644, doi:10.1038/s41586-021-03207-w (2021).

34 Tre-Hardy, M. et al. Immunogenicity of mRNA-1273 COVID vaccine after 6 months surveillance in health care workers; a third dose is necessary. J Infect 83, 559–564, doi:10.1016/j.jinf.2021.08.031 (2021).

35 Tre-Hardy, M., Cupaiolo, R., Wilmet, A., Beukinga, I. & Blairon, L. Waning antibodies in SARS-CoV-2 naive vaccinees: Results of a three-month interim analysis of ongoing immunogenicity and efficacy surveillance of the mRNA-1273 vaccine in healthcare workers. J Infect 83, 381–412, doi:10.1016/j.jinf.2021.06.017 (2021).

36 Abbasi, J. SARS-CoV-2 Variant Antibodies Wane 6 Months After Vaccination. JAMA 326, 901–901, doi:10.1001/jama.2021.15115 (2021).

37 Levin, E. G. et al. Waning Immune Humoral Response to BNT162b2 Covid-19 Vaccine over 6 Months. N Engl J Med 385, e84, doi:10.1056/NEJMoa2114583 (2021).

38 Sterlin, D., Malaussena, A. & Gorochov, G. [IgA dominates the early neutralizing antibody response to SARS-CoV-2 virus]. Med Sci (Paris) 37, 968–970, doi:10.1051/medsci/2021154 (2021).

39 Wang, Z. et al. Enhanced SARS-CoV-2 neutralization by dimeric IgA. Sci Transl Med 13, doi:10.1126/scitranslmed.abf1555 (2021).

40 Kusnadi, A. et al. Severely ill COVID-19 patients display impaired exhaustion features in SARS-CoV-2-reactive CD8(+) T cells. Sci Immunol 6, doi:10.1126/sciimmunol.abe4782 (2021).

41 Boppana, S. et al. SARS-CoV-2-specific circulating T follicular helper cells correlate with neutralizing antibodies and increase during early convalescence. PLoS Pathog 17, e1009761, doi:10.1371/journal.ppat.1009761 (2021).

42 Gong, F. et al. Peripheral CD4+ T cell subsets and antibody response in COVID-19 convalescent individuals. J Clin Invest 130, 6588–6599, doi:10.1172/JCI141054 (2020).

43 Rha, M. S. et al. PD-1-Expressing SARS-CoV-2-Specific CD8(+) T Cells Are Not Exhausted, but Functional in Patients with COVID-19. Immunity 54, 44–52 e43, doi:10.1016/j.immuni.2020.12.002 (2021).

44 Liao, M. et al. Single-cell landscape of bronchoalveolar immune cells in patients with COVID-19. Nat Med 26, 842–844, doi:10.1038/s41591-020-0901-9 (2020).

45 Grau-Exposito, J. et al. Peripheral and lung resident memory T cell responses against SARS-CoV-2. Nat Commun 12, 3010, doi:10.1038/s41467-021-23333-3 (2021).

46 Wu, T. et al. Lung-resident memory CD8 T cells (TRM) are indispensable for optimal cross-protection against pulmonary virus infection. J Leukoc Biol 95, 215–224, doi:10.1189/jlb.0313180 (2014).

47 Channappanavar, R., Zhao, J. & Perlman, S. T cell-mediated immune response to respiratory coronaviruses. Immunol Res 59, 118–128, doi:10.1007/s12026-014-8534-z (2014).

48 Sekine, T. et al. Robust T Cell Immunity in Convalescent Individuals with Asymptomatic or Mild COVID-19. Cell 183, 158–168 e114, doi:10.1016/j.cell.2020.08.017 (2020).

49 Oja, A. E. et al. Trigger-happy resident memory CD4(+) T cells inhabit the human lungs. Mucosal Immunol 11, 654–667, doi:10.1038/mi.2017.94 (2018).

50 Goliwas, K. F. et al. Methods to Evaluate Cell Growth, Viability, and Response to Treatment in a Tissue Engineered Breast Cancer Model. Sci Rep 7, 14167, doi:10.1038/s41598-017-14326-8 (2017).

51 Goliwas, K. F., Miller, L. M., Marshall, L. E., Berry, J. L. & Frost, A. R. Preparation and Analysis of In Vitro Three Dimensional Breast Carcinoma Surrogates. J Vis Exp, doi:10.3791/54004 (2016).

52 Goliwas, K. F. et al. Extracellular Vesicle Mediated Tumor-Stromal Crosstalk Within an Engineered Lung Cancer Model. Front Oncol 11, 654922, doi:10.3389/fonc.2021.654922 (2021).

53 Naba, A., Clauser, K. R. & Hynes, R. O. Enrichment of Extracellular Matrix Proteins from Tissues and Digestion into Peptides for Mass Spectrometry Analysis. J Vis Exp, e53057, doi:10.3791/53057 (2015).

