## Extended Figure Legends for "Local SARS-CoV-2 Peptide-Specific Immune Responses in Lungs of Convalescent and Uninfected Human Subjects"

**Extended Figure 1:** *Ex Vivo* Maintenance of Lung Tissue over 14 Days Culture. **A**. Histologic architecture of human lung tissue is maintained following 14 days *ex vivo* culture. **B**. Lactate dehydrogenase (LDH) levels are stable during the culture period, indicating no increase in cell death during culture. **C-H**. Frequency of cell populations within the lung, comparing starting tissue to tissues cultured *ex vivo* for 7 or 14 days. **I**. The proportion of extracellular matrix (ECM) proteins comparing starting tissue to tissues cultured for 14 days. n=3-11, statistical differences were evaluated using one-way ANOVA with Sidak’s multiple comparisons testing.

**Extended Table 1:** Patient Demographics.

**Extended Table 2:** COVID-19 Convalescent Extended Demographics.

**Extended Figure 2:** Epithelial Cell Population Characterization following SARS-CoV-2 Peptide Exposure. **A**. Gating strategy for epithelial cells. **B**. Proportion of epithelial cell populations in starting tissues of UN and COVID19^+^ lung tissues. **C**. Baseline differences in epithelial cell populations within UN and COVID19^+^ lung tissues. **D**. Changes in epithelial cell populations over the culture period (day 0 vs. day 6 control). **E.** Impact of SARS-CoV-2 peptide exposure on epithelial cell populations in UN and COVID19^+^ lung tissue. n=8 UN and n=4 COVID19^+^. Statistics shown in blue are comparisons between control and peptide exposed samples within each group (UN and COVID19^+^). Statistics shown in black are the change in response between UN and COVID19^+^ for each peptide when compared to the corresponding controls.

**Extended Figure 3:** Endothelial Cell Characterization with SARS-CoV-2 Peptide Exposure. **A**. Gating strategy for endothelial cells. **B**. Baseline differences in endothelial cells within UN and COVID19^+^ lung tissues. **C**. Changes in endothelial cells over the culture period (day 0 vs. day 6 control). **D.** Impact of SARS-CoV-2 peptides on endothelial cells in UN and COVID19^+^ lung tissue. n=8 UN and n=4 COVID19^+^. Statistics shown in blue are comparisons between control and peptide exposed samples within each group (UN and COVID19^+^). Statistics shown in black are the change in response between UN and COVID19^+^ for each peptide when compared to the corresponding controls.

**Extended Figure 4:** Impact of SARS-CoV-2 Peptide Exposure on TNF-α producing immune cells. **A**. Baseline differences in TNF-α^+^ immune cells within UN and COVID19^+^ lung tissues. **B**. Changes in TNF-α^+^ immune cells over the culture period (day 0 vs. day 6 control). **D.** Impact of SARS-CoV-2 peptides on TNF-α^+^ immune cells in UN and COVID19^+^ lung tissue. n=8 UN and n=4 COVID19^+^.

**Extended Figure 5:** Proliferating, Antigen-specific, and Interferon-γ Producing CD4^+^ T cell changes in response to SARS-CoV-2 Peptide Exposure. **A**. Gating strategy for CD4^+^ T cells. **B-E**. Baseline differences in proliferating (**B**), antigen-specific (**C**), and interferon-γ producing (**D-E**) CD4^+^ T cell populations within UN and COVID19^+^ lung tissues. **D**. Changes in epithelial cell populations over the culture period (day 0 vs. day 6 control). **F-I.** Impact of SARS-CoV-2 peptide exposure on proliferating (**F**), antigen-specific (**G**), and interferon-γ producing (**H-I**) CD4^+^ T cell populations in UN and COVID19^+^ lung tissue. n=8 UN and n=4 COVID19^+^. Statistics shown in blue are comparisons between control and peptide exposed samples within each group (UN and COVID19^+^). Statistics shown in black are the change in response between UN and COVID19^+^ for each peptide when compared to the corresponding controls.

**Extended Figure 6:** Proliferating, Antigen-specific, and Interferon-γ Producing CD8^+^ T cell changes in response to SARS-CoV-2 Peptide Exposure. **A**. Gating strategy for CD8^+^ T cells. **B-E**. Baseline differences in proliferating (**B**), antigen-specific (**C**), and interferon-γ producing (**D-E**) CD8^+^ T cell populations within UN and COVID19^+^ lung tissues. **D**. Changes in epithelial cell populations over the culture period (day 0 vs. day 6 control). **F-I.** Impact of SARS-CoV-2 peptide exposure on proliferating (**F**), antigen-specific (**G**), and interferon-γ producing (**H-I**) CD8^+^ T cell populations in UN and COVID19^+^ lung tissue. n=8 UN and n=4 COVID19^+^. Statistics shown in blue are comparisons between control and peptide exposed samples within each group (UN and COVID19^+^). Statistics shown in black are the change in response between UN and COVID19^+^ for each peptide when compared to the corresponding controls.

**Extended Figure 7:** T Cell Population Changes over the Culture Period. **A-O.** Changes in T cell populations over the culture period (day 0 vs. day 6 control). n=8 UN and n=4 COVID19^+^. Statistics shown in blue are comparisons between control and peptide exposed samples within each group (UN and COVID19^+^). Statistics shown in black are the change in response between UN and COVID19^+^ for each peptide when compared to the corresponding controls.

**Extended Figure 8:** Gating Strategy for Memory T cells, Starting Populations, and Changes over the Culture Period. **A**. Gating strategy for Memory T cells. **B-C**. Proportion of memory CD4+ (**B**) and CD8+ (**C**) T cell populations in starting tissues of UN and COVID19^+^ lung tissues. **D-M**. Changes in memory T cell populations over the culture period (day 0 vs. day 6 control). n=8 UN and n=4 COVID19^+^. Statistics shown in blue are comparisons between control and peptide exposed samples within each group (UN and COVID19^+^). Statistics shown in black are the change in response between UN and COVID19^+^ for each peptide when compared to the corresponding controls.

**Extended Figure 9:** Gating Strategy for B cells, Starting Populations, and Changes over the Culture Period. **A**. Gating strategy for B cells. **B**. Proportion of B cell populations in starting tissues of UN and COVID19^+^ lung tissues. **C-F**. Changes in immature B cells (**C**), memory B cells (**D**), class-switched memory B cells (**E**), and transitional B cells (**F**) in starting UN and COVID19+ tissues. G-J. Changes in immature B cells (**G**), memory B cells (**H**), class-switched memory B cells (**I**), and transitional B cells (**J**) in UN and COVID19+ tissues with peptide treatment. **K-Q**. Changes in B cell populations over the culture period (day 0 vs. day 6 control). n=4 UN and n=3 COVID19^+^. Statistics shown in blue are comparisons between control and peptide exposed samples within each group (UN and COVID19^+^). Statistics shown in black are the change in response between UN and COVID19^+^ for each peptide when compared to the corresponding controls.

**Extended Figure 10:** CD138^+^ plasma cells within COVID-19^+^ samples secrete IgA in response to peptide stimulation. **A**. Photomicrographs of UN sample #3 showing CD138^+^ plasma cells (red) and IgA (green) in control tissue and in response to N peptide addition. **B**. Photomicrographs of COVID-19^+^ samples #5 and #8 showing CD138^+^ plasma cells (red) and IgA (green) in control tissue and in response to M, N, or S peptide addition. White arrows pointing to CD138^+^ plasma cells secreting IgA, grey dashed arrows pointing to CD138^+^ plasma cells without IgA secretion.
