## Extended Figures for "Local SARS-CoV-2 Peptide-Specific Immune Responses in Lungs of Convalescent and Uninfected Human Subjects"

**Extended Figure 1**:

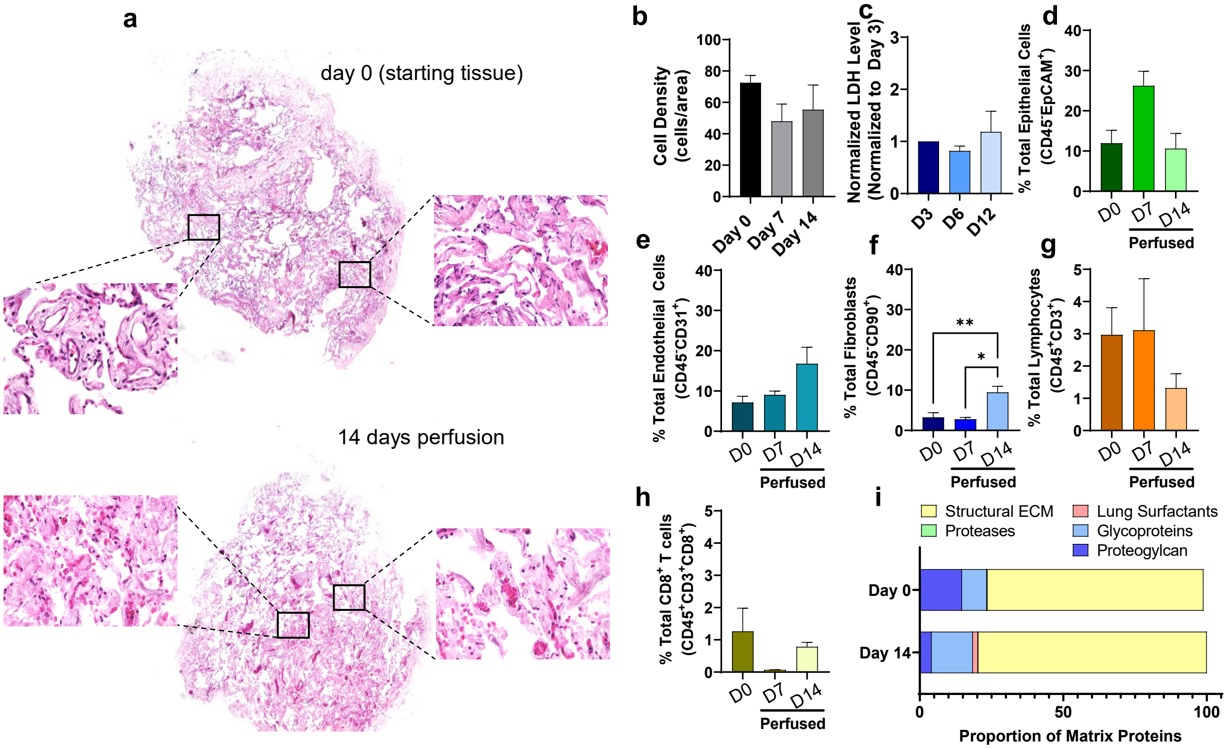

#### Extended Table 1

| | **Uninfected** | **COVID-19+** |
|---|---|---|
| **Tissues collected** | 8/2020-4/2021 | 10/2020-4/2021 |
| **Number collected** | 8 | 4 |
| **Median Age (Range)** | 65 (48-76) | 62 (46-73) |
| **Male** | 3/8 (37.5 %) | 2/4 (50%) |
| **Ethnicity** | | |
| ***Caucasian*** | 7/8 (87.5%) | 2/4 (50%) |
| ***Black*** | 1/8 (12.5%) | 2/4 (50%) |
| ***Average Convalescence Period (Range)*** | -------- | ~ 71 Days (28-104 days)* |

* One patient tested positive twice before surgery (2 and 9 months before surgical resection) the most recent positive test was used to calculate the convalescence period

**Extended Table 2**

| COVID-19⁺ Samples | | | | |
|---|---|---|---|---|
| **Sample** | **Convalescence Period Before Resection** | **Sex** | **Vaccination Status** | **Other** |
| 5 | 28 d | Female | Not vaccinated | |
| 8 | 77 d | Male | Not vaccinated | |
| 10 | 104 d | Female | Vaccinated | First dose only; 27 days before resection |
| 11 | 75 d | Male | Not vaccinated | Tested positive twice (~2 and 9 months prior to resection) |

### Extended Figure 2

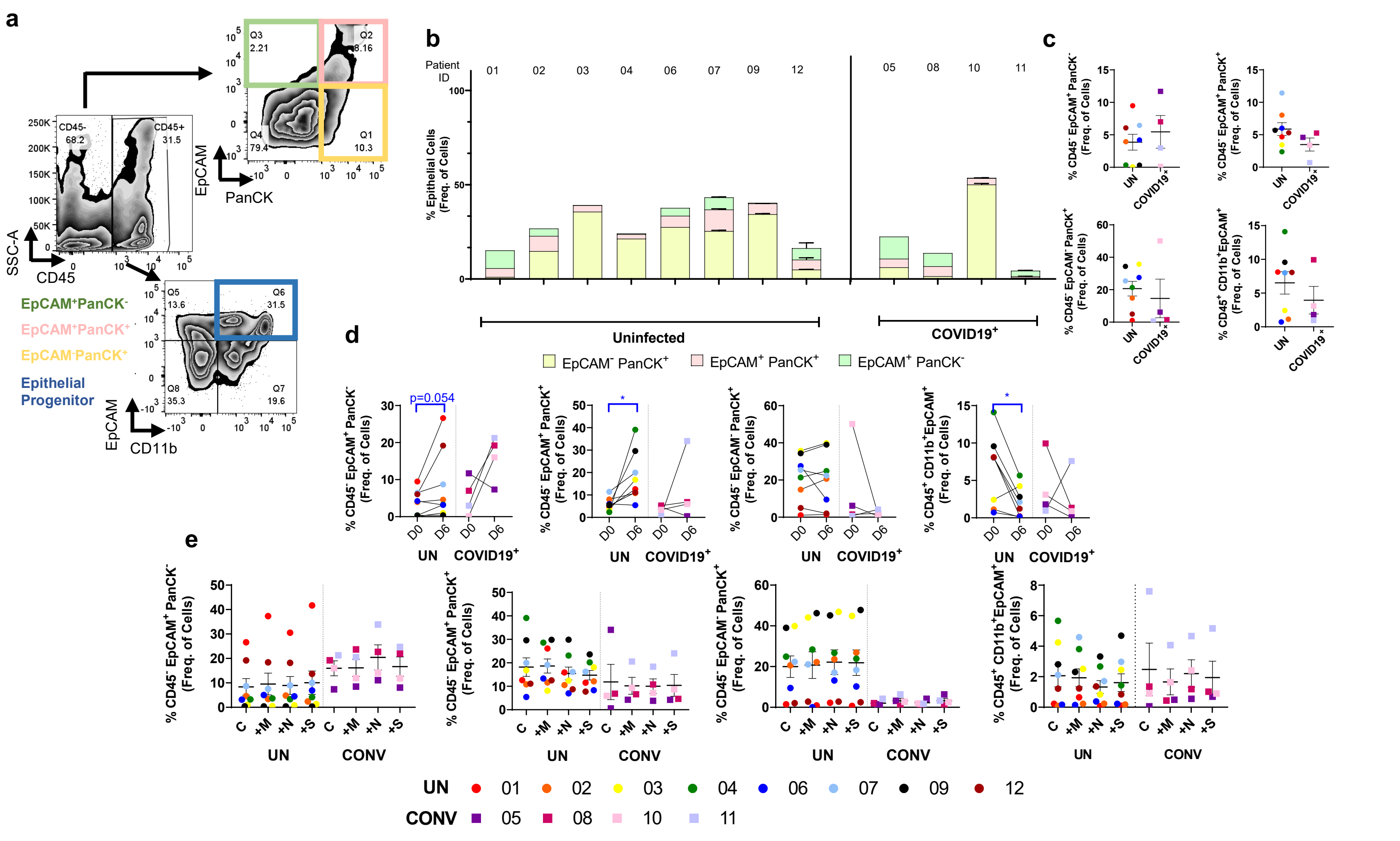

### Extended Figure 3

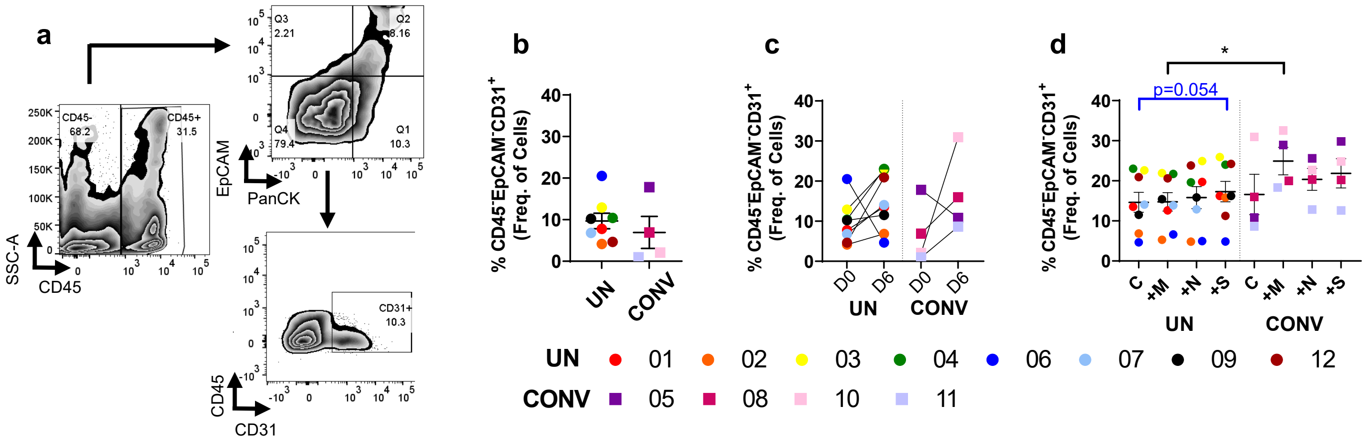

#### Extended Figure 4

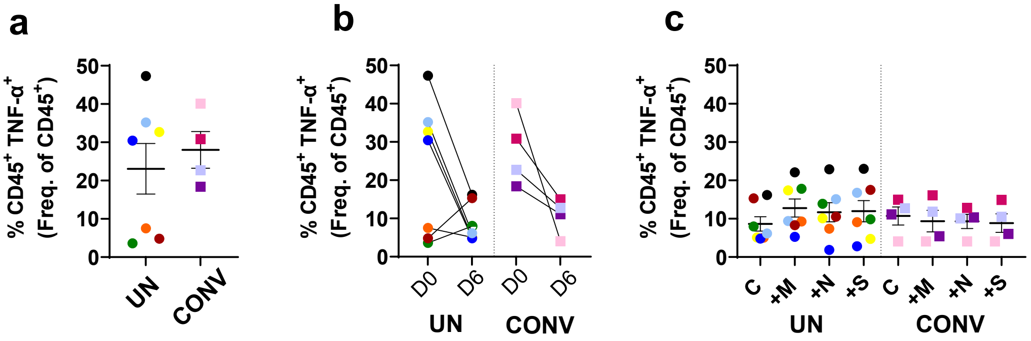

**Extended Figure 5**

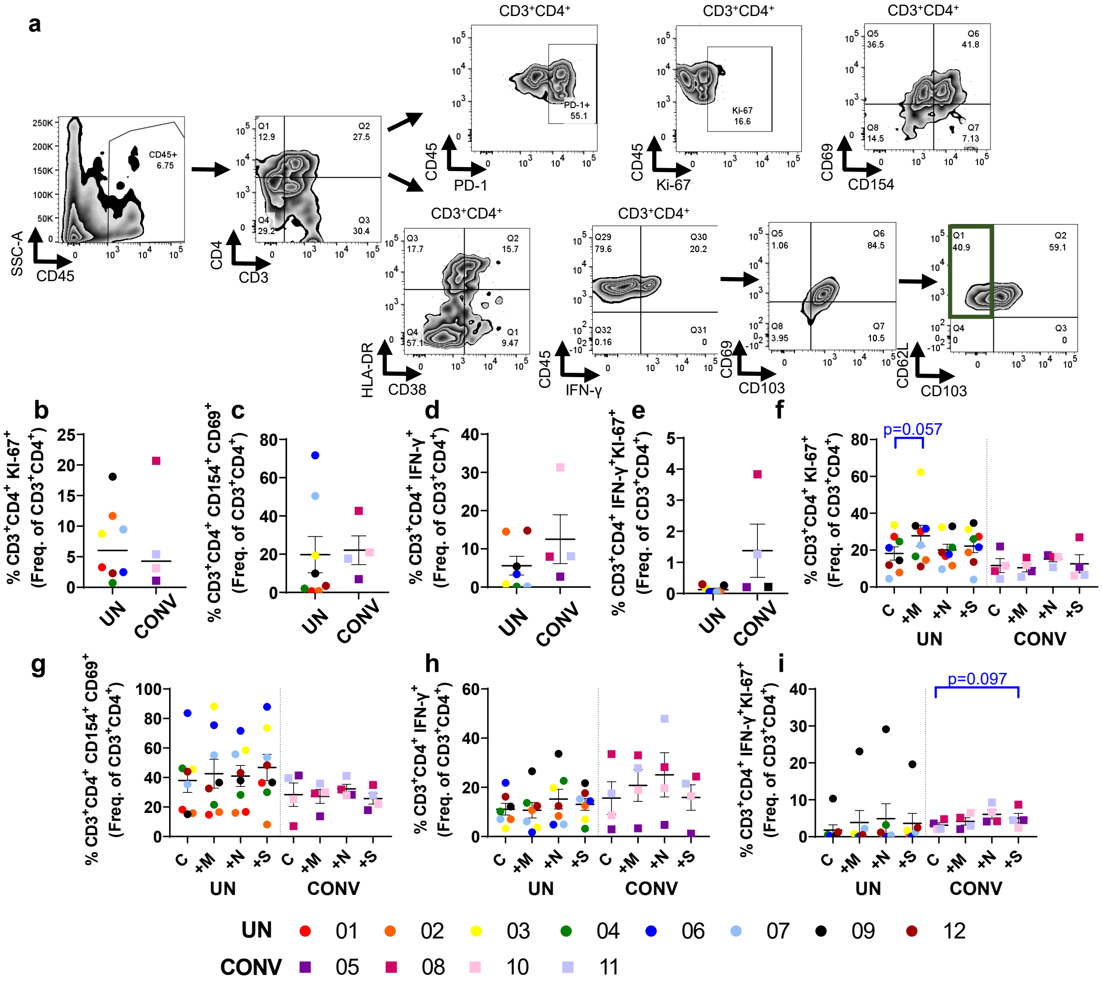

### Extended Figure 6

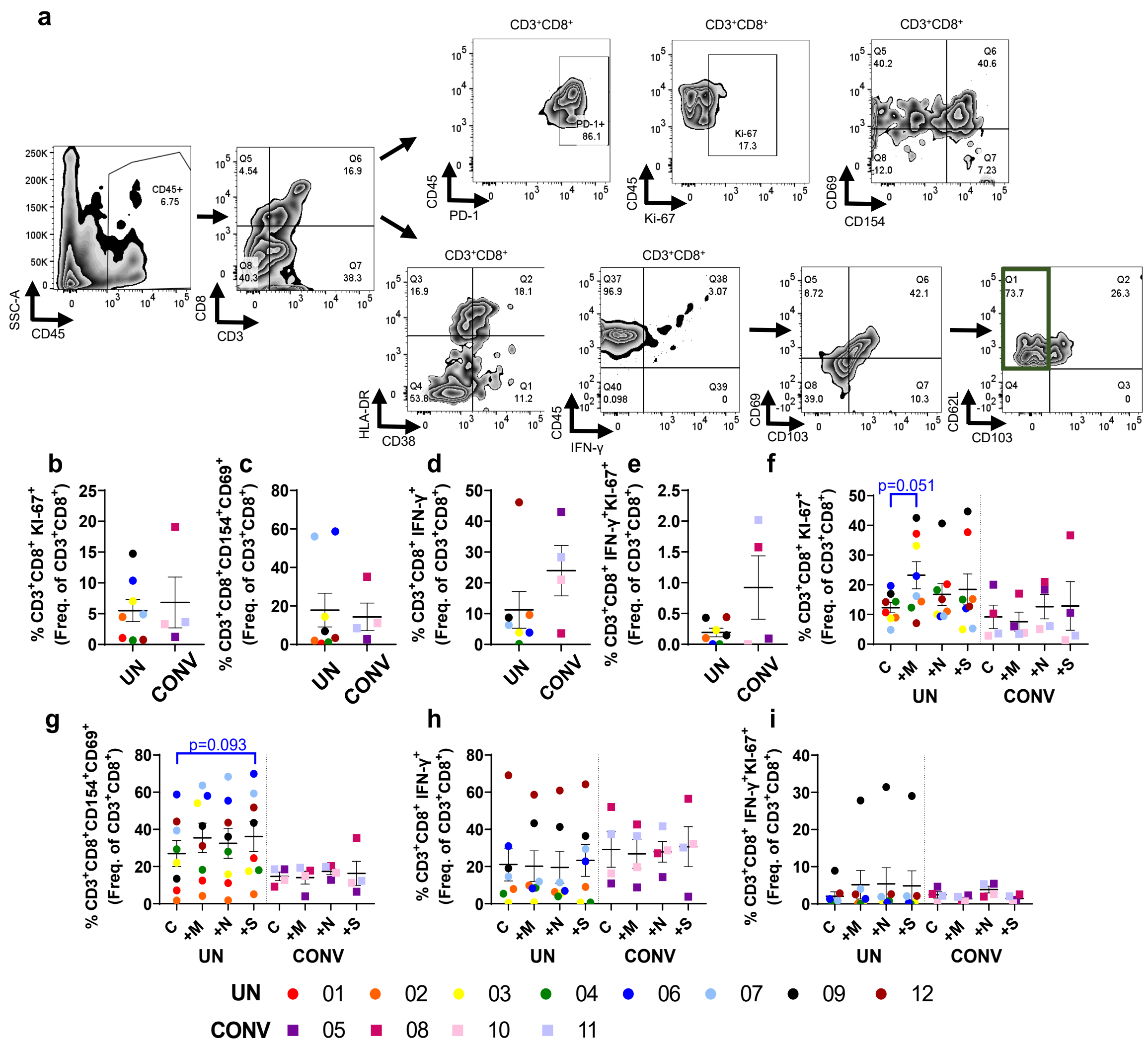

#### Extended Figure 7

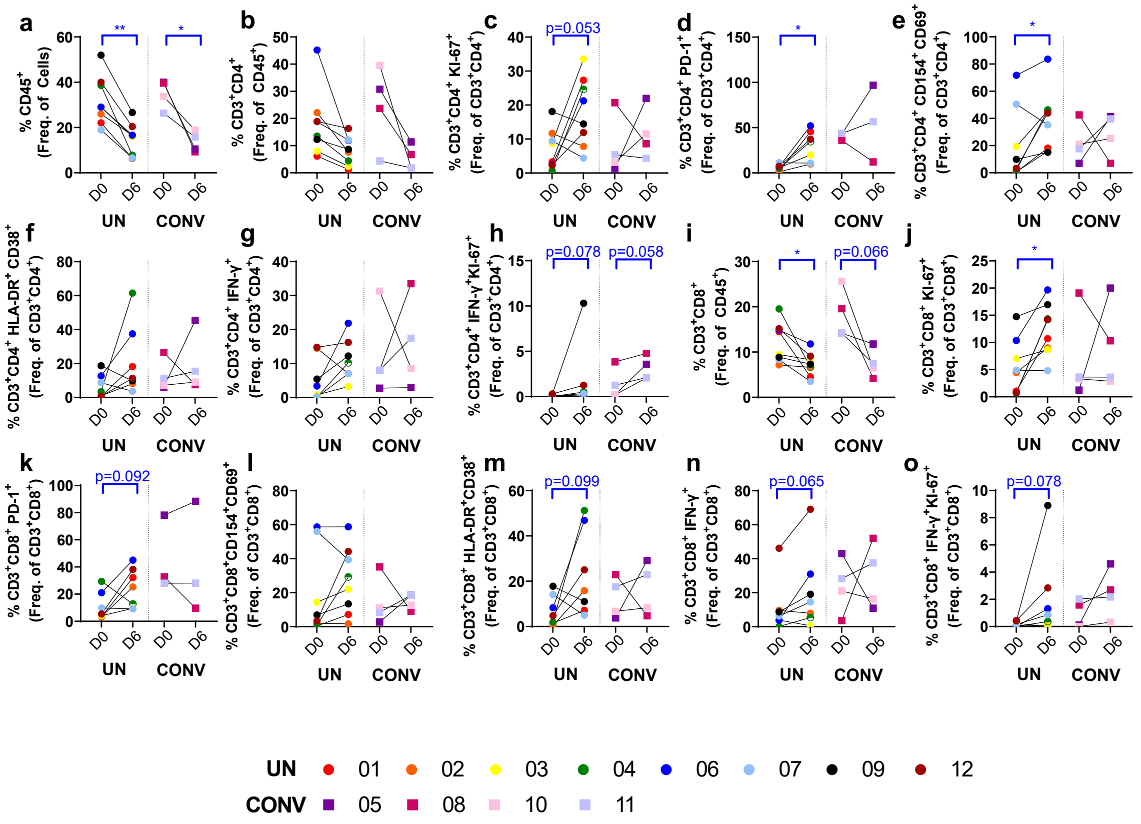

### Extended Figure 8

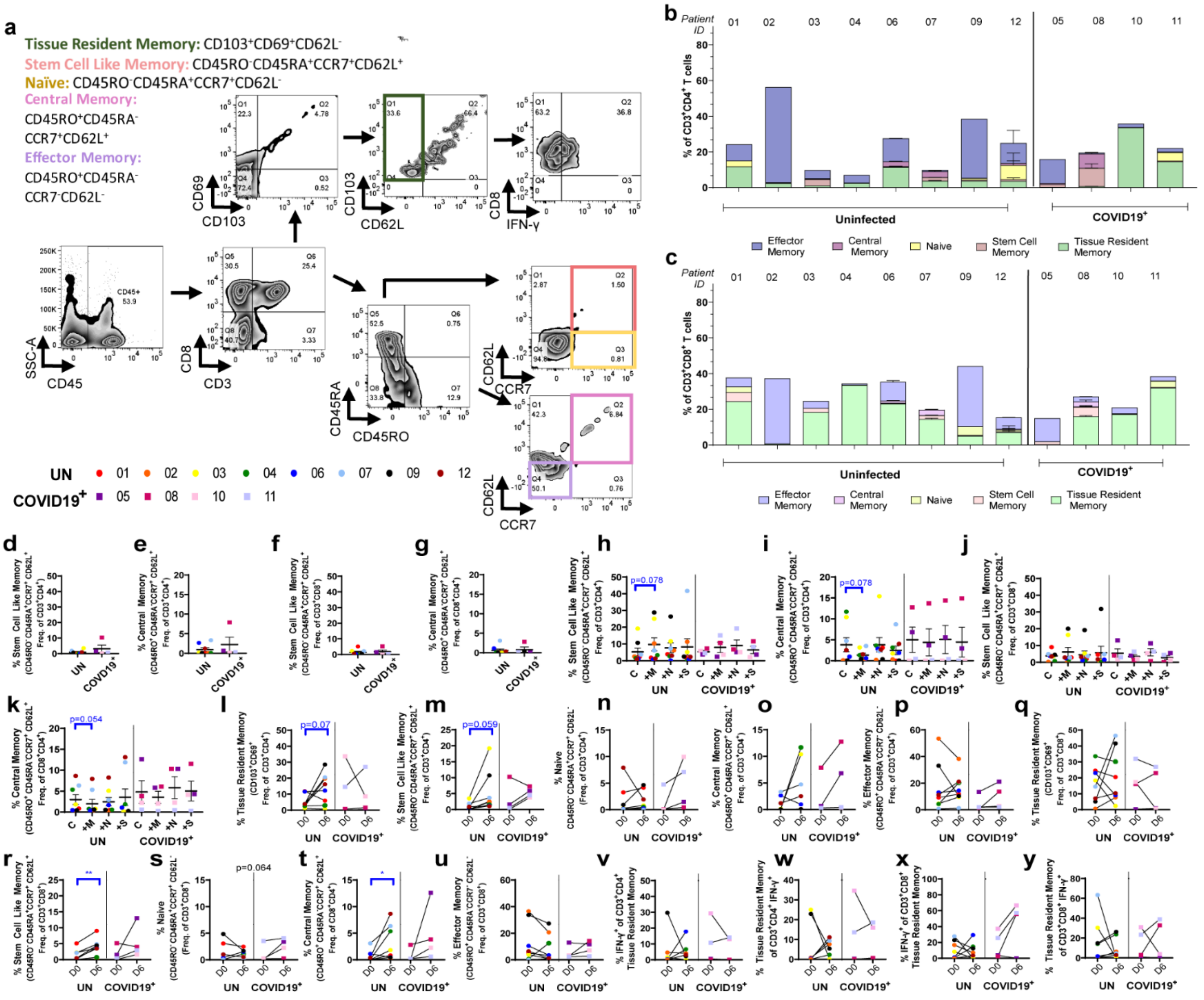

### Extended Figure 9

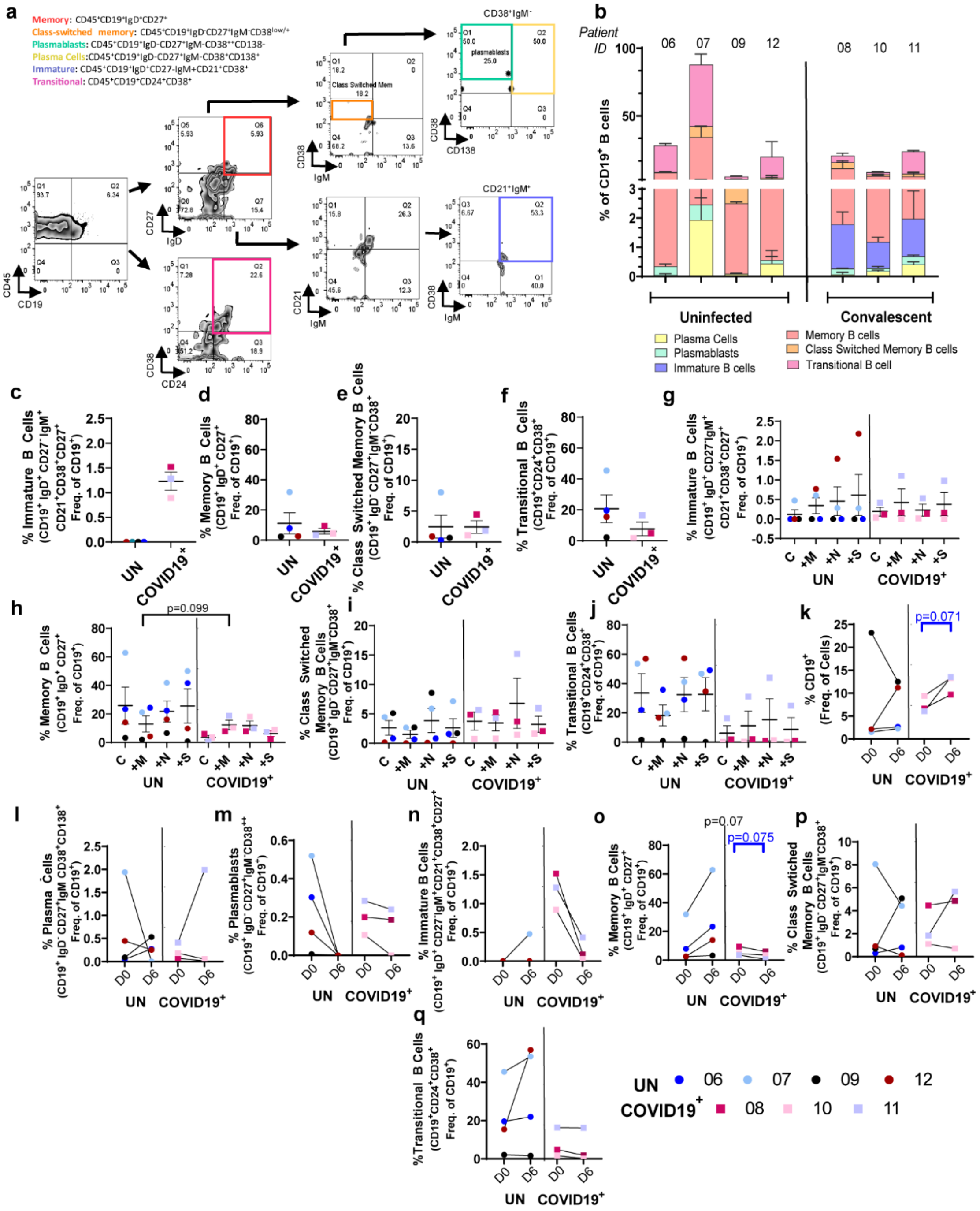

**Extended Figure 10**

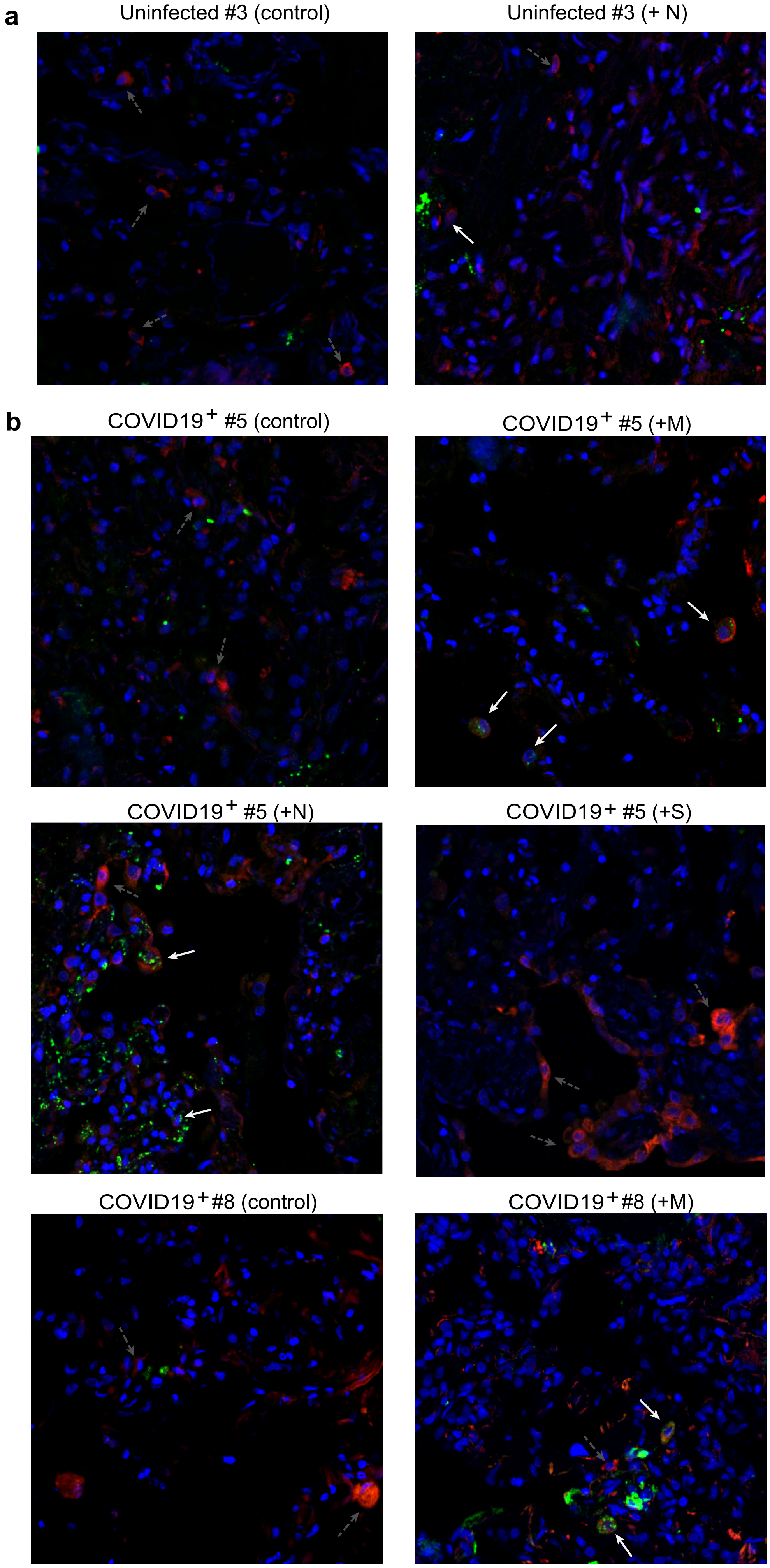
